# Quantification of a Viromed Klinik Akut V 500 disinfection device to reduce the indirect risk of SARS-CoV-2 infection by aerosol particles

**DOI:** 10.1101/2020.10.23.20218099

**Authors:** Christian J. Kähler, Thomas Fuchs, Rainer Hain

## Abstract

Indoor SARS-CoV-2 infections by droplets and aerosols are currently considered to be particularly significant. FFP2/3 respirator masks, which fit tightly and gap free, generally provide very good protection. In public transport, while shopping or in waiting rooms, they are therefore ideally suited to protect against direct and indirect infection. Unfortunately, these masks make it difficult to breathe and can be uncomfortable to wear in the long run. Therefore, these masks should be worn for a maximum of 3 × 75 minutes per day. These masks are therefore hardly suitable for schools or at work. The question therefore arises as to how people in closed rooms can be permanently protected from a SARS-CoV-2 infection. Large safety distances provide both self protection and protection of third parties, but they do not protect against indirect infection if the virus load in the room is high. Mouth and nose covers only offer protection of others against direct infection, but they do not protect the user against indirect infection. The same applies to faceshields and small protective walls. Indirect infections can be effectively prevented by free ventilation with windows or air conditioning systems that supply 100% outside air into the room, provided the air exchange rate is at minimum six times the room volume per hour. However, free ventilation by means of windows is rarely efficient enough, and in winter at the latest, it is no longer possible to open windows without wasting massive amounts of energy and endangering the health and well-being of people. The operation of air conditioning systems is also very energy-intensive during the cold season. Furthermore, most buildings do not have air conditioning systems. The question is therefore, how a largely safe protection against an indirect SARS-CoV-2 infection can be realized in closed rooms without wasting thermal energy and thus valuable resources. Technically, the problem can be solved with mobile disinfection devices or room air cleaners that separate the dangerous aerosol particles or inactivate the viruses by UV radiation or by contact with charge carriers. The potential of these devices is great and, since many German manufacturers produce these devices, they are also available. However, many of the devices offered do not provide effective protection because the volume flow is too small, the separation efficiency of the filters is too low and the performance of the UV and ionization unit is too weak. The Viromed Klinik Akut V 500 disinfection unit appears to meet the performance requirements and therefore the device is analyzed and evaluated in this study for its suitability to protect against SARS-CoV-2 infection.

## 1. Introduction

According to the current state of research, SARS-CoV-2 is mainly transmitted via droplets and aerosol particles that are produced when breathing, speaking, singing, coughing, or sneezing and are inhaled and exhaled through the air we breathe [1, 2, 3, 4]. Direct infection, in which many emitted droplets and aerosol particles are inhaled over a short distance (less than 1.5 m) by an uninfected person, can be effectively prevented with the aid of particle-filtering respirator masks (FFP2/3 or better), since these respirator masks reliably separate droplets and aerosol particles during inhalation and exhalation up to a specified size class, if they are tightly and closely attached to the face [5, 6]. If these masks are used without an outlet valve, large safety distances between persons are not required to prevent direct infection. In addition, nothing needs to be done to prevent indirect infections caused by an increased viral load in the room, since particle-filtering respiratory masks also provide reliable protection against this transmission route [5, 7].

Particle-filtering respiratory masks are used for occupational safety in hospitals, laboratories, isolation wards, operating theatres and many technical work areas where fine dust and substances harmful to health are handled (e.g. grinding, welding, soldering). The argument that the SARS-CoV-2 viruses cannot be reliably separated because the viruses are smaller than 0.16 μm is not correct, because SARS-CoV-2 is transported by droplets or droplet nuclei and these are significantly larger than individual viruses and can be reliably separated by suitable particle-filtering masks [6]. It should also be noted that very small aerosol particles often do not carry viruses and even if they do carry a virus, many of these very small aerosol particles would have to be inhaled to cause an infection [8]. It is estimated that a dose of at least 500 − 2000 viruses is required to cause a SARS-CoV-2 infection [9, 10].

A major disadvantage of particle filtering masks without a valve is that they make breathing difficult. In order to avoid overstraining the wearer, they should only be worn for a maximum of 75 minutes at a time [11]. A break of 30 minutes is recommended before the masks are worn again [11]. Particle-filtering masks are therefore very suitable for use on public transport in the city or when shopping, as these activities can often be completed in less than 75 minutes and because this “personal isolation” does not restrict mobility [7]. In school it is already becoming more difficult to meet these time requirements and in the office, with an 8-hour working day, other protective measures must be taken, as the masks should not be worn for more than 3 × 75 minutes per day [11]. However, it must also be taken into account that these masks not only make breathing more difficult and are sometimes uncomfortable, but also cause considerable costs in the long run. If a school class with 25 children uses FFP2/3 respirators daily for a unit price of 4 Euros, then 200 school days per year would result in a total of 20000 Euros per class and year or 800 Euros per child per year. In addition to these costs, it must also be taken into account that the masks generate waste and thus cause further costs. Therefore, this solution is neither economically nor ecologically sensible.

Alternatively, people in public buildings with public access or communal areas could maintain sufficiently large safety distances to avoid direct infection. In schools, offices or waiting rooms, however, this option is not feasible either, as the corresponding rooms are not available. Doubling the distance between students in both directions would require a quadrupling of the required classroom space. The shortage of classrooms could be solved by running the school in four shifts, but there are not enough teachers, and hiring them would involve immense costs and would therefore not be financially viable. Apart from the costs, the teaching staff is not available, so this option is not feasible. Furthermore, this concept has to take into account that distances alone in a closed room cannot guarantee safety from a SARS-CoV-2 infection. Since the viral load in a room depends on the number of infected persons and their length of stay and activity, additional measures must be taken to limit the viral load in the room air, as otherwise indirect infections may occur. In order to enable a communal use of the room over several hours, it is recommended to limit the direct danger of infection either by simple mouth-nose covers, face shields or small transparent walls between people sitting at a table (school desk) and the indirect danger of infection by other measures.

One way to prevent indirect infections is to use free ventilation through open windows, so that the virus load in the room cannot reach critical levels. However, free ventilation only works physically if there is either a difference in temperature between inside and outside or if the wind blows in front of the windows [12, 13]. A temperature difference often does not exist and if it does exist, it is quickly reduced during free ventilation, so this mechanism is usually not efficient or is only effective for a short time. The wind in front of the window is also rarely strong enough to ensure adequate ventilation. Since the effectiveness of free ventilation depends on factors that cannot be influenced (temperature, wind speed, size/position of windows), the question is how to ventilate when these physical mechanisms cannot be used. However, it must also be taken into account that free ventilation during the cold season leads to colds and impairs the well-being of people. Furthermore, manual ventilation must be considered and one must want to and be able to do it (in many schools the windows cannot be opened). Another very important argument against free ventilation is the waste of thermal energy. Houses are being elaborately and cost-intensively insulated in order to contribute to limiting global warming. The demand of some people that the climate targets should be of secondary importance during the pandemic is incomprehensible. Instead of wasting thermal energy via free ventilation, measures should therefore be taken to reconcile the protection of people during the pandemic with climate goals.

Many buildings are equipped with modern air conditioning systems, which ensure that contaminated air is removed in a controlled manner and filtered or “fresh” outside air is added from outside. The main advantage of HVAC systems compared to free ventilation is that they continuously provide adequate indoor air quality and regular manual regulation by means of windows is not necessary. In order to reduce the indirect SARS-CoV-2 infection risk, however, the systems must be operated correctly. For energy reasons, they are often operated with low fresh air supply and simple filters. To prevent indirect SARS-CoV-2 infections, however, a large proportion of fresh air (preferably 100%) or very good filtering of the room air with class H13 / H14 filters is required [14]. Existing air handling systems are usually operated with simple class F7 / F9 filters and not with high-quality class H13 / H14 filters, which are capable of reliably separating 99.995% of aerosol particles from a size of 0.1 − 0.3 μm. As a result, many installed air handling units do not allow for an energetically favourable and safe recirculation air operation. A retrofit of the air handling units is usually not feasible either, since the installation of filters of class H13 / H14 would lead to a reduction of the volume flow due to the increased pressure resistance. However, due to the hazardous nature of SARS-CoV-2, air exchange rates of at least 6 per hour must be required. Estimates which are supposed to justify that an air exchange rate of 1 − 3 per hour offers sufficient protection against indirect infection are based on false assumptions. It should be remembered that in rooms where infectious persons are treated, air exchange rates of 12 − 15 are recommended or prescribed [15, 16, 17]. Therefore, it is not understandable why the emission of dangerous viruses in schools and offices should be counteracted with an air exchange rate of 1 − 3. An air exchange rate of 6 per hour can be regarded as a good compromise between technical feasibility and safety of a SARS-CoV-2 infection and thus a compromise between cost and benefit. If high performance HVAC systems are available, they should be operated in such a way that the air exchange rate is at least 6 per hour and the proportion of fresh air should be 100%. Although this mode of operation is energetically poor, the disadvantages of free ventilation can at least be avoided. However, it must be taken into account that many buildings do not have air handling systems and therefore the question arises how normal operation / stay can be realized in public buildings. Furthermore, the question is whether the energetic disadvantages of air conditioning systems cannot be eliminated by a recirculation mode, in which the room air is passed through a filter that separates virus-contaminated aerosol particles or inactivates the viruses with UV-C or by ionizing the room air in order to prevent infection. The air in the room would then not have to be heated or the humidity adjusted, as would be necessary with air handling systems that feed a large proportion of outside air into the room. This would reduce costs and save valuable resources.

A simple and proven method of separating droplets and aerosols from the room air is offered by room air cleaners [8]. If they are equipped with a filter of the class H13 / H14 and the volume flow is large enough to lead six times the amount of the room volume per hour through the filter, then these devices are basically suitable to prevent indirect SARS-CoV-2 infections. Furthermore, these devices also separate pollen and fine dust particles, so that allergy sufferers and people with respiratory diseases benefit from the devices. Besides the pure separation of droplets and aerosol particles, other physical mechanisms can be used to inactivate the viruses so that they can no longer cause infections. In principle, this inactivation can be realized by means of UV radiation. However, if only UV technology is used, it is usually not possible to realize large volume flows, because at large volume flows the residence time of the viruses in the electromagnetic radiation is too short or the distance to the UV source becomes too large to ensure reliable inactivation of the viruses. The problem could be solved by increasing the UV radiation output, but then harmful ozone can be produced. UV technology should therefore only be used in combination with other technologies to combat viruses at high volume flows.

Another technology that can be used to combat viruses is based on the ionization of air and aerosol particles. The electrically charged air can directly inactivate the viruses on contact with them, destroying their ability to cause infection. The harmful effect of ionized air on viruses and other pathogens is used in hospitals, for example to accelerate the healing of chronic wounds [18, 19]. In addition, charged ions accumulate on aerosol particles and electrostatic interaction may subsequently cause them to combine with other aerosol particles to form larger clusters. These clusters can be filtered better than single particles. However, it has to be considered that in normal living rooms the aerosol concentration in the room is usually very low, so that a cluster formation is quite unlikely because the distances between the aerosol particles are too large. Consequently, we believe that direct inactivation of the viruses by ionized air plays a more important role in preventing SARS-CoV-2 infections than clustering.

Within the scope of this study, the Viromed Klinik Akut V 500 disinfection device is analyzed, which uses all three physical mechanisms to combat the viruses and germs in indoor air simultaneously. Due to the complexity of the flow problem, an experimental approach is used for the analysis, since only in this way can the many influencing factors be physically correctly recorded and evaluated.

According to the manufacturer, the Viromed Klinik Akut V 500 disinfection device has the following features [20]:

1. volume flow up to a maximum of 330 m^3^/h.
2. fine filter on the suction side of the unit to separate dust particles etc. from the room air.
3. ionization of the air molecules by means of cold plasma to destroy/inactivate the reproductive ability of viruses by means of electrical charges.
4. electrostatic separation of charged aerosol particles and germs.
5. UV-C radiation unit for destruction of viral RNA by means of electromagnetic radiation.
6. ionization of air molecules for destruction of viruses by means of electric charges in the room and separation by cluster formation.

## 2. Experimental setup and performance of the PIV experiments

The aim of the first test series was to quantify the flow field in the vicinity of the sterilizer at a maximum volume flow of 330 m^3^/h. For this purpose, the average air flow velocity and the turbulent air movement at the inlet and outlet of the sterilizer must be quantitatively determined. Both variables are important, as people easily perceive air movements as disturbing. According to DIN 1946 Part 2 [21], flow velocities of less than 0.3 m/s can be assumed to be non-disturbing when the unit is not in operation. Since the air movement felt is composed of the average and turbulent flow movement, the sum of both components must be less than 0.3 m/s on average.

Particle Image Velocimetry (PIV) was used to determine these two quantities in spatial resolution [22]. In this measurement technique, the position of artificially generated aerosol particles in a laser light section is registered with digital cameras at two points in time and then the local displacement of the particle images is determined using digital image processing methods. From the displacement of the particle images, the spatially resolved velocity distribution in the measuring plane can then be determined, taking into account the time interval between the measurements and a calibration factor. Figure 1 shows pictures of the test setup (left) and during a PIV measurement (right).

**Figure 1:**
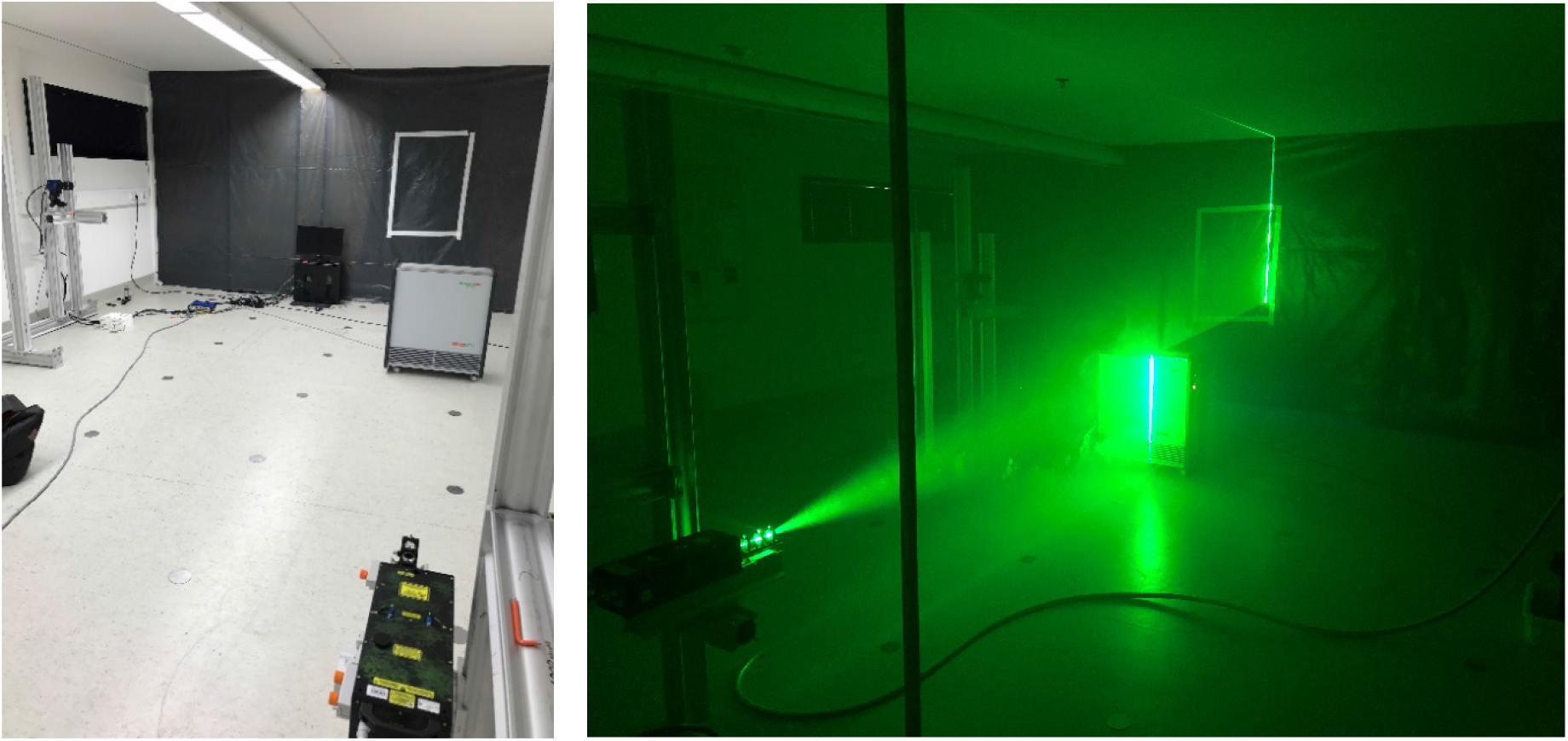
Experimental setup with disinfection unit, a double pulse laser and a sCMOS camera (left) and recording during measurement with PIV (right)

For the experiments a PCO.edge 5.5 sCMOS camera with a Zeiss Milvus lens with a focal length of 50 mm was used. The aerosol particles were generated from di-2-ethylhexyl sebacate (DEHS) with a seeding generator from the company PIVTEC. The mean diameter of the aerosol particles is 1 μm and the size distribution ranges from 0.1 − 2 μm [23]. A Quantel Evergreen 200 laser was used to illuminate the particles and the beam was fanned out into a light sheet using various lenses [22]. The measuring system was controlled by the software DaVis from LaVision GmbH, which was also used for data evaluation. The PIV equipment was traversed to record the entire flow field in front of and above the sterilizer.

## 3. Air movement in the vicinity of the Viromed Klinik Akut V 500 disinfection device

The following two figures show the results of the PIV measurements at a maximum volume flow of the Viromed Klinik Akut V 500 disinfection unit of approx. 330 m^3^/h. In figure 2 the amount of velocity is visualized in color-coded form. The direction of the mean air movement can be recognized by the orientation of the vector arrows and the length of the vectors illustrates the velocity. Figure 3 shows the magnitude of the mean turbulent air movement as a color-coded gradient. The larger the turbulent flow movement is, the more the air movement fluctuates around the value of the mean local flow velocity. The vectors symbolize the magnitude and direction of the mean flow velocity according to figure 2 for orientation.

**Figure 2:**
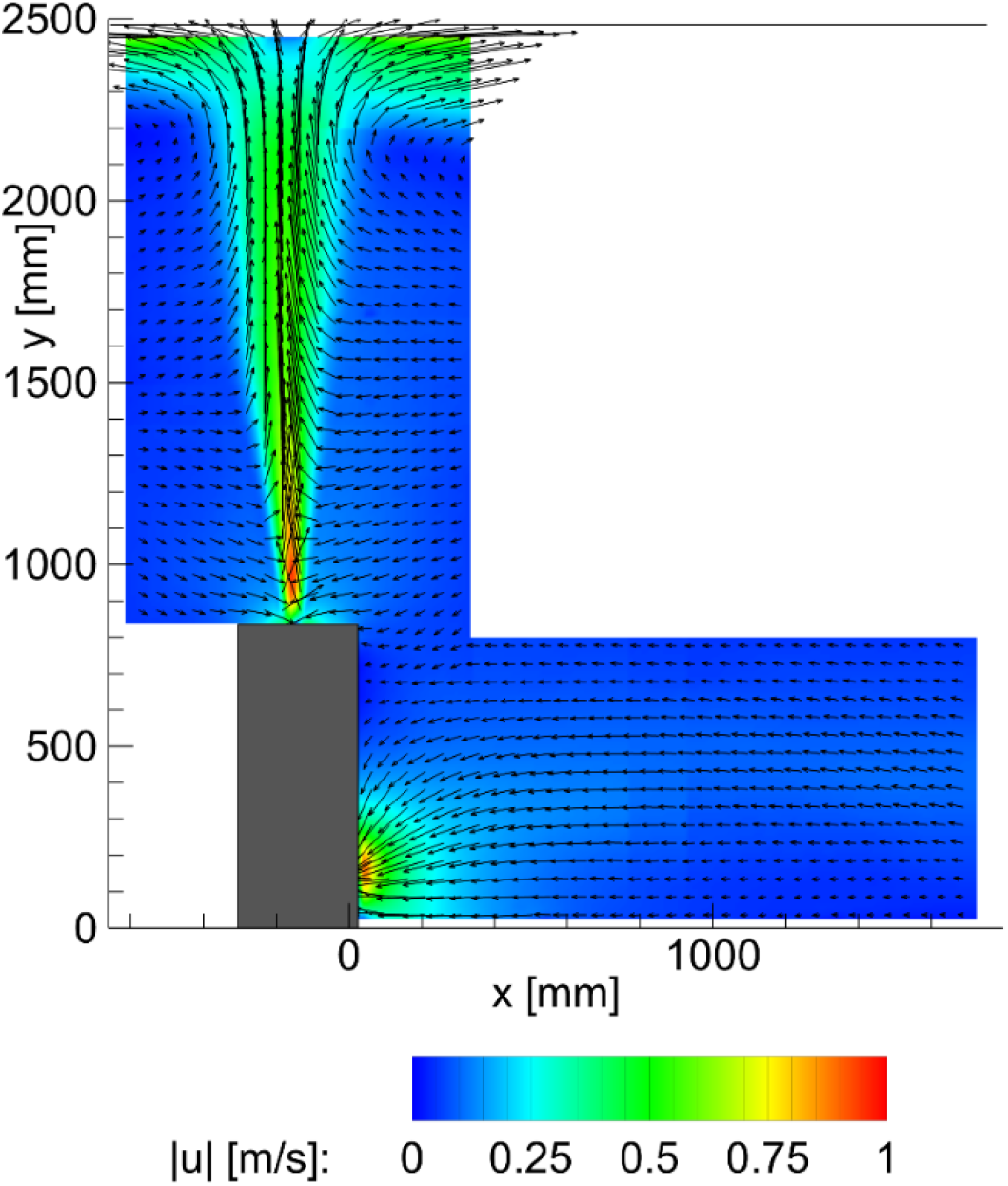
Magnitude and direction of the mean flow movement for 330 m^3^/h

**Figure 3:**
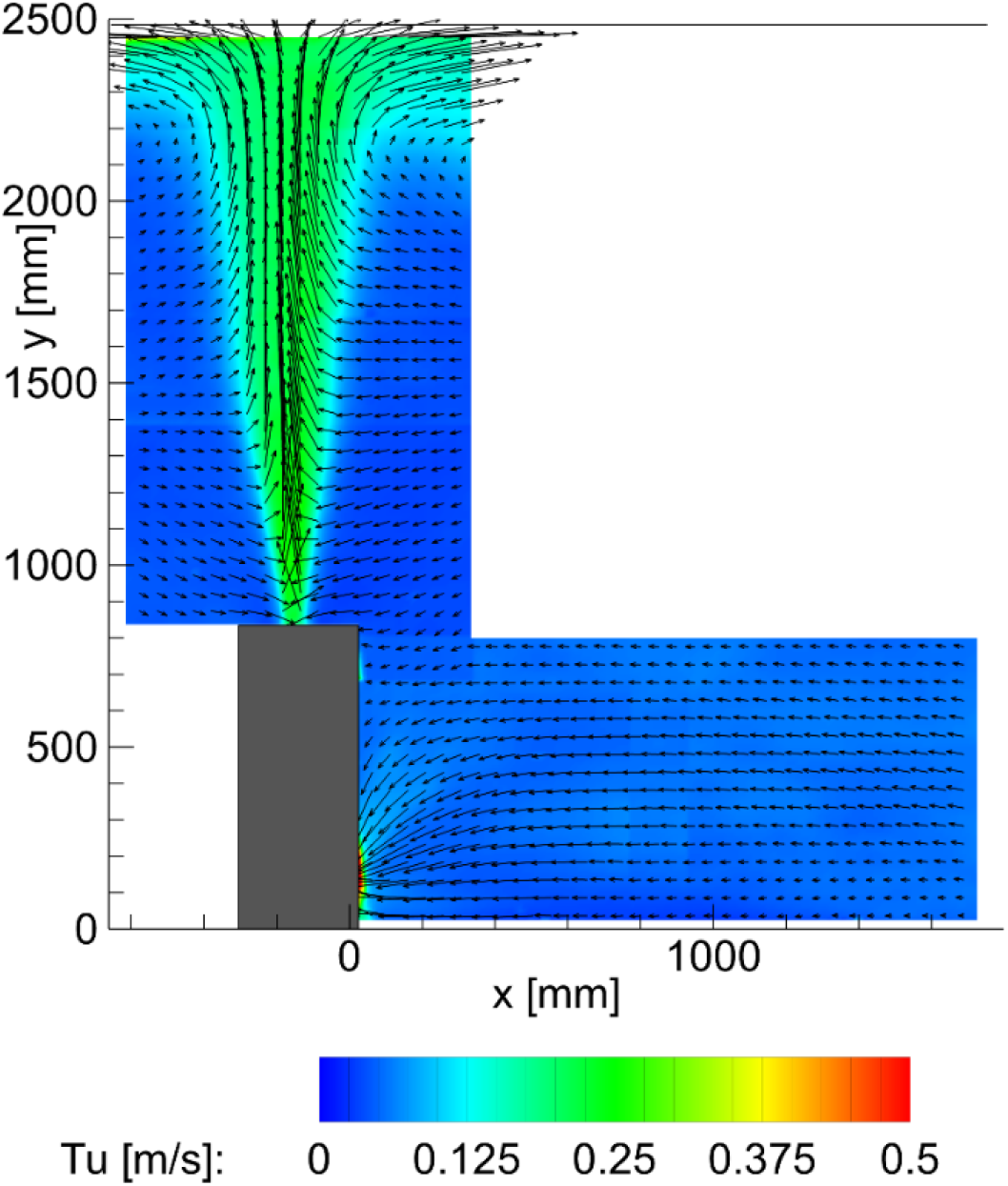
Turbulent air movement averaged over time at 330 m^3^/h

The quantitative PIV measurement results clearly show that the sum of the mean flow velocity and the mean turbulent motion in the environment of the instrument does not exceed the recommended limits according to DIN 1946 part 2. Therefore, even if people stay in the immediate vicinity of the sterilizer, no impairment of the people’s well-being by uniform or fluctuating air movements is to be expected. With a maximum volume flow of 330 m^3^/h, flow velocities of up to 0.5 m/s are only achieved up to a distance of about 0.2 m in front of the intake area. However, these only occur in the foot area, so that they cannot be perceived as disturbing by persons. In the sensitive head and body area, the average flow velocities and the superimposed turbulent air movement are significantly less than 0.3 m/s in total.

Higher flow velocities are achieved directly above the air outlet. However, since the air flows vertically upwards out of the device, the air flow cannot impair the well-being. Furthermore, due to the vertical outlet of the free jet, blocking of the flow by objects in the room is not easily possible. Blocking of the outlet jet could lead to an impairment of the filter performance and should therefore be avoided. As an interim result of this analysis, it can be stated that the operation of the Viromed Klinik Akut V 500 disinfection unit keeps the air movement within the recommended limits.

Figure 2 shows very clearly that the outflowing air flows up to the ceiling and is then deflected in both directions in approximately equal parts. The air then spreads along the ceiling due to the Coandă effect until it is deflected downwards by the walls of the room and then flows back to the intake area of the unit. This air movement is shown schematically by arrows in Figure 4 (left). In order for a large-scale circulation movement to develop in the room, the spread of the air flow should not be blocked by larger objects in the ceiling area (long protruding lamps, ceiling projections, ceiling beams, …) [8, 24]. The more undisturbed the air can propagate, the more efficiently and quickly the contaminated room air is passed through the unit. If the flow propagation is disturbed, more complex room air flows are formed, as shown in Figure 4 (right) as an example. This room air movement can lead to a reduced filter performance or reduced inactivation rate of the viruses.

**Figure 4:**
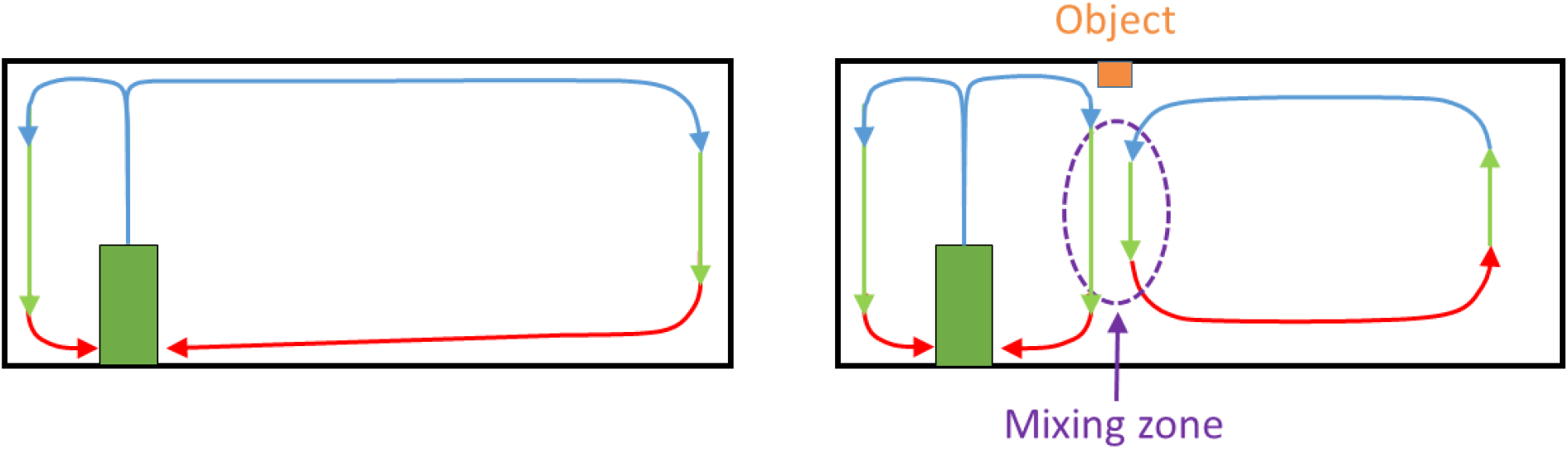
Idealized representation of the air flow in an empty room (left) and in the presence of a bar projecting into the room (right). In reality, the flow phenomena are three-dimensional.

In order to compensate for this undesired air movement, a slightly larger volume flow can be set at the disinfection unit. However, as the operating costs increase with increasing volume flow and the noise level rises, care should be taken to position the unit appropriately in the room. In order to enable an energetically efficient and effective filtering of the room air, it is recommended to position the sterilizing unit in the middle of the longest side of the room and the air jet hitting the ceiling should be able to spread along the ceiling without being blocked.

If the vectors in Figure 2 are examined closely, it can be seen that parts of the air returning from the room interact directly with the free jet exiting the unit and propagate back into the room without being guided through the unit. This is a completely natural flow-mechanical process, which is called entrainment. If the volume flow of the unit is sufficiently large, it is ensured that the air volume passing the unit is filtered in a sufficiently short time and that the viruses are separated or inactivated. This is ultimately ensured by the turbulent mixing in the room. However, the goal should be to keep the time for filtering the room air as short as possible so that an air exchange rate of 6 per hour can be achieved in an energetically favorable way. How fast the filtering of the room air and thus the inactivation of the viruses finally takes place can be determined by concentration measurements.

## 4. Test setup and execution of the concentration measurements

In order to quantitatively determine the filter performance of the disinfection device and thus the separation and inactivation of viruses, the temporal decrease in particle concentration was first measured simultaneously at three positions in a 80 m^2^ room. Since the room volume is 200 m^3^, but the unit only has a volume flow of 330 m^3^/h, an air exchange rate of 6 per hour cannot be achieved. Therefore several units would have to be operated simultaneously in the room. However, the decay curves allow the results to be transferred to other room sizes. Smaller rooms are analyzed in the next chapter.

Figure 5 shows the geometry and dimensions of the room in top view. Since the room is too large for a single unit of this power class according to the manufacturer’s specifications, experiments were carried out with two units placed at positions A1 and A2. The two units were then installed at the corner positions B1 and B2 in order to quantify the influence of the installation site on the filter performance and the time until the viruses were inactivated. Finally, for comparison purposes, measurements were performed with a single device located at position B1.

**Figure 5:**
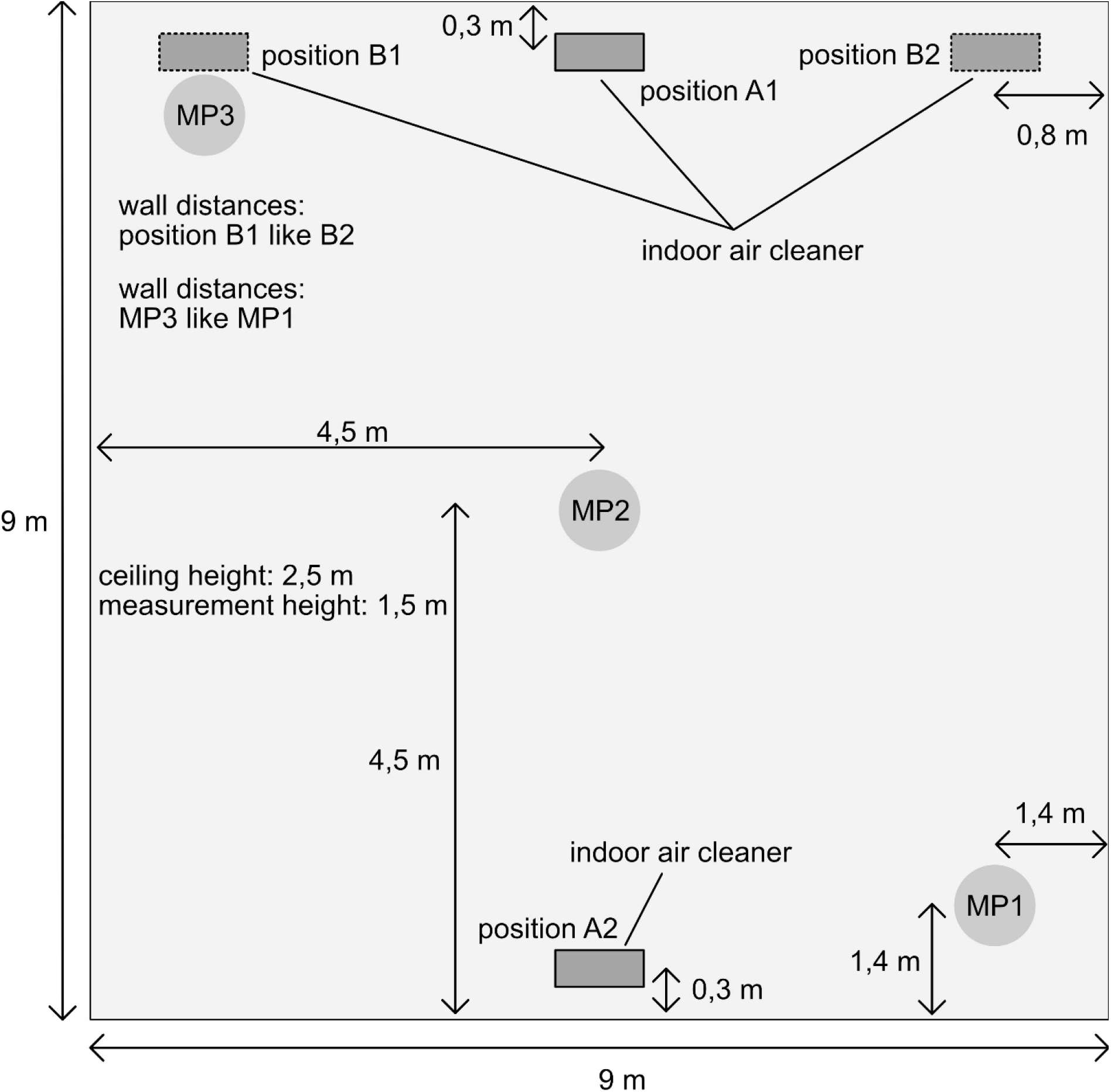
Arrangement of the components in the room for concentration measurements

The measurement positions are marked MP1 − MP3 in Figure 5. The particle imaging method was used to determine the temporal decrease of aerosol particles in the ambient air. In this method, the room is first nebulized homogeneously and with high concentration with very long-lived aerosol particles whose size distribution corresponds to the aerosol particles emitted when breathing, speaking, and singing. The longevity of the aerosol is very important, as otherwise a systematic falsification of the measurement results by evaporation would occur. Furthermore, the small size of the aerosol particles is important because large aerosol particles would sink over time, which would also cause systematic measurement errors. The aerosol particles are illuminated with a pulsed laser and imaged with a camera with a suitable lens and stored digitally for further processing. The number of particle images on the sensor corresponds to the number of aerosol particles in the illuminated measuring volume. The number of particle images on the sensor must not be too large, because overlapping particle images would systematically falsify the count. For this reason, the imaging scale of the optical system and the initial concentration of the aerosol must be selected sensibly. For the detection of the particle images, digital filters are applied which suppress the background noise. As a result of this image preprocessing only the images of the aerosol particles remain on the image, which are then automatically counted. Without this image preprocessing, stochastic image noise could be misinterpreted as a signal, which would lead to systematic measurement errors. By taking images at a fixed frequency over a sufficiently long period of time, the individual particle images can be reliably counted in each individual image. The result of the measurements is the number of aerosol particles in the measurement volume as a function of time.

From the temporal course of the particle number, important parameters such as the decay constant, the half-life and the residence time of the aerosol particles in space can be determined. The value of the decay constant theoretically corresponds exactly to the air exchange rate. The half-life is a measure of how long it takes until the number of aerosol particles at the measurement position under consideration is reduced by 50%. After twice the half-life time, the concentration of the aerosol particles has consequently dropped to 25% of the initial value. The residence time is the average time the released aerosol particles need to travel from the respective measuring position to the deposition site in the sterilizer. On the way from the release site to the deposition site, the viral load will naturally decrease due to the convective movement and turbulent mixing of the room air, so that the risk of infection is reduced with increasing distance from the source due to the flow.

In order to be able to analyze the functionality of the Viromed Klinik Akut V 500 disinfection unit in a 80 m^2^ room, the aerosol concentration was measured simultaneously at 3 independent positions diagonally in the room. Using a PIVTEC seeding generator, aerosol particles with a size distribution between 0.1 − 2 μm and a mean diameter of about 1 μm were generated from di-2-ethylhexyl sebacate (DEHS) [19]. To illuminate the aerosol particles, the output beam of a Quantel Evergreen 200 laser was aligned diagonally across the room. To capture images of the aerosol particles in the measuring volumes (MP1 − MP3), 3 PCO.edge 5.5 sCMOS cameras with Zeiss Milvus lenses with a focal length of 50 mm were used. The individual cameras and the laser were controlled by LaVision’s DaVis software so that the recordings of all cameras were performed synchronously. The acquisition rate for the measurements was 1 Hz. Figure 6 shows in a distorted panoramic image the arrangement of the used components in space.

**Figure 6:**
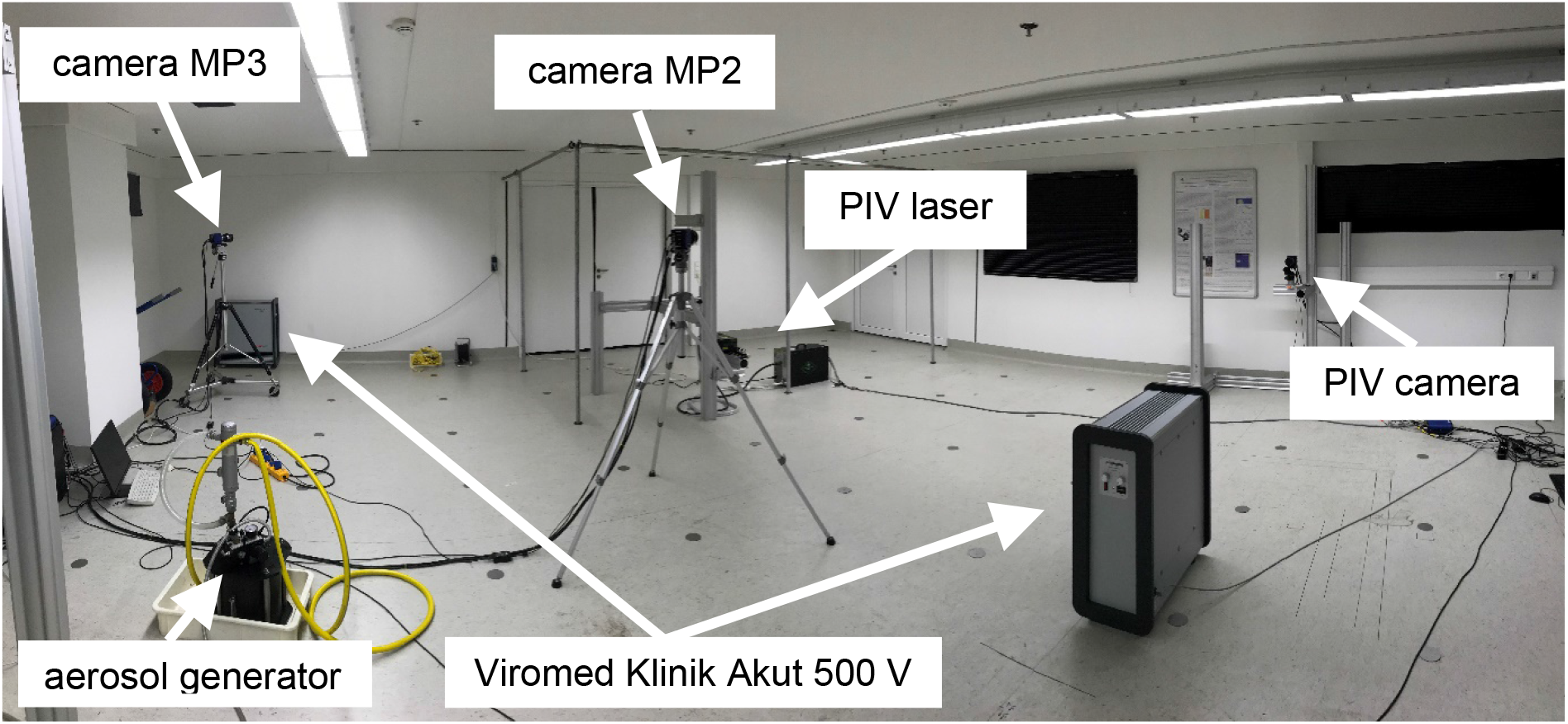
Optically distorted panoramic image of the experimental room with the components for PIV and concentration measurements

## 5. Concentration measurements in a square room with 80 m^2^

Using the PIV experimental setup mentioned in section 2, the separation efficiency at the outlet of the disinfection unit was first visually analyzed. For this purpose, the entire room was homogeneously nebulized with DEHS aerosol particles with a diameter of 0.1 − 2 μm and a mean diameter of approx. 1 μm [23]. With the help of a laser light sheet in the outflow area it was examined whether aerosol particles are still coming out of the outlet of the device. Figure 7 shows that the environment of the free jet is completely contaminated with aerosol particles (white areas). In contrast, the free jet emerging centrally from the device is free of aerosol particles (dark area). The fact that the filtered air does not appear completely black in the illustration is due to the efficient mixing with the surrounding room air, which is clearly visible in Figure 7. The process by which contaminated air areas enter the clean jet is called entrainment [12]. Analyses in which aerosol particles were only introduced in the intake area clearly show that they are reliably separated by electrostatic filtration.

**Figure 7:**
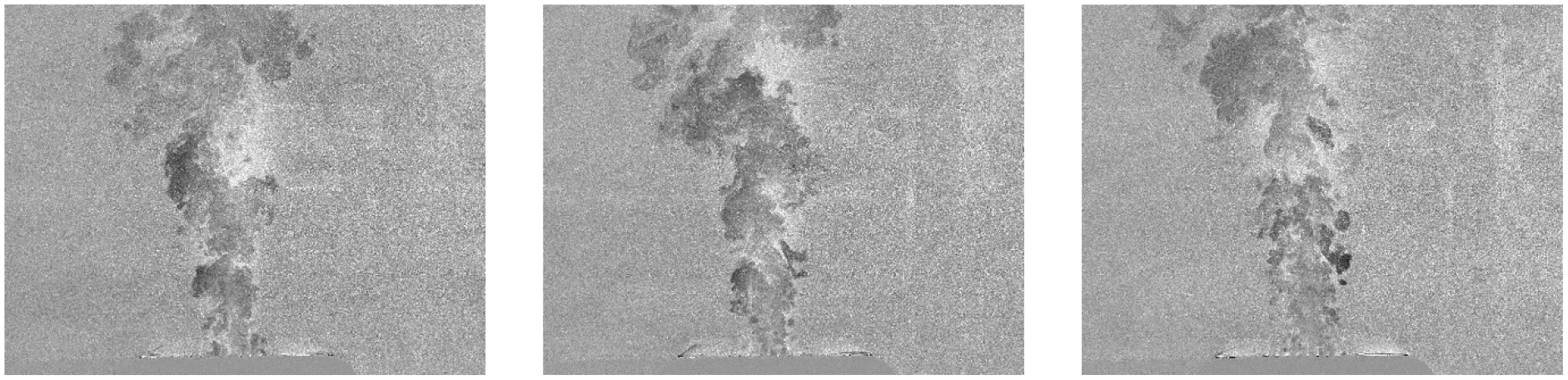
Representation of the aerosol distribution in the outflow area at a volume flow rate of 330 m^3^/h

In order to be able to quantitatively evaluate the filter performance and the inactivation of the viruses in the 80 m^2^ room, Figure 8 (left) shows the course of the measured aerosol particle number as a function of time. In addition, the temporal decrease of the aerosol concentration when the disinfection unit is switched off is shown as a reference. Since all openings in the room were sealed airtight and the very small aerosol particles hardly settle at all, the particle concentration in the reference measurement decreases only very slowly. The result of the reference measurement also shows that small long-lived droplet nuclei or droplets remain in the air for hours under conditions in which the evaporation rate is in equilibrium with the condensation rate. It is therefore important to technically limit the virus load in the room so that, if possible, no infectious virus concentration can form in the room.

**Figure 8:**
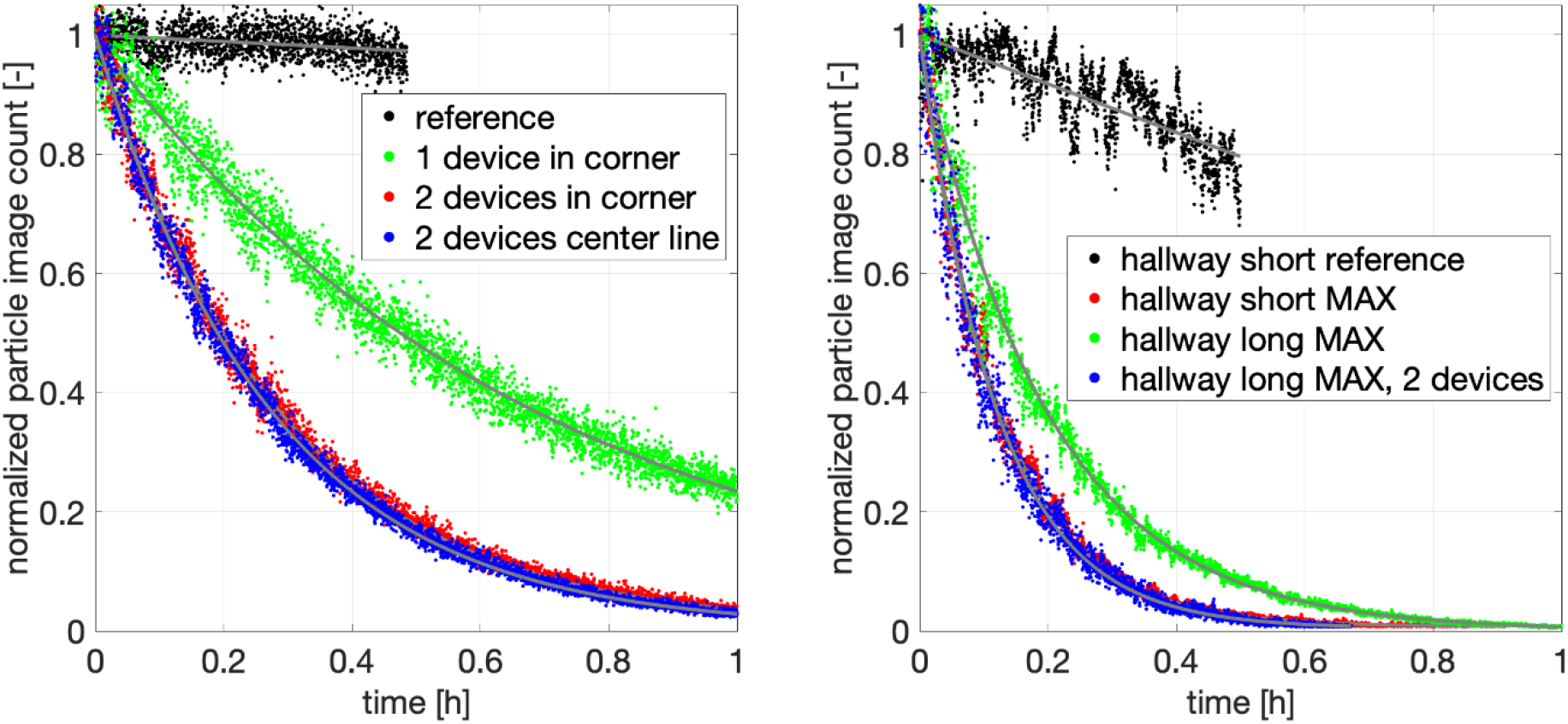
Decrease of aerosol concentration over time for different volume flow rates and associated exponential fit functions

It is also obvious that aerosol particles can be transported by the air flow over very long distances (in principle many kilometers) if the spread is not prevented by walls. However, it must be taken into account that in this process the concentration and thus the probability of infection is reduced very quickly due to two processes. On the one hand, turbulent diffusion causes a spatial dispersion of the aerosol, which also occurs when the mean flow velocity is zero. On the other hand, the aerosol released over a period of time is spatially strongly stretched and consequently diluted if the mean flow velocity is not zero. If, for example, 1 liter of air is exhaled over a period of 2 seconds during light physical exertion and the surrounding air flows past the point of exit at an average speed of 10 m/s, the exhaled air is stretched over a range of 20 m due to the flow. The concentration will therefore decrease mathematically by a factor of 200 and thus the viral load in the wake of the person from whom the aerosol is exhaled. If turbulent diffusion is also taken into account, the concentration will decrease again significantly. These flow-mechanical processes explain why outside closed rooms a SARS-CoV-2 infection is very unlikely and therefore could hardly be detected. Furthermore, not all aerosol particles carry viruses [8]. In the open air, therefore, an aerosol infection is extremely unlikely if there is sufficient wind speed or movement of persons, provided that distances between persons of at least 1.5 m are maintained.

The comparison of the different measurement curves in Figure 8 (left) shows the decrease of the aerosol concentration in the 80 m^2^ room as a function of time for the different configurations. The comparison illustrates the effect of the positioning and number of devices on the filter performance and thus on the reduction of the viruses in the room air. The effect of the volume flow rate is clearly visible. A doubling of the number of units and thus of the volume flow rate halves the time required to obtain a certain concentration. If the aerosol concentration in a room with a certain volume is to be halved in a certain time, then these measured curves can be used to estimate how large the volume flow must be to achieve the desired target.

The exponentially decreasing course of the aerosol particle number allows to quantitatively determine characteristic quantities, which are essential for the evaluation of the efficiency of the disinfection device. The decay constant, which results from the exponential decrease of the concentration in figure 8, is a measure for the efficiency of the filtration. The larger the value, the faster the decrease and the better the filtering effect and the shorter the time required to filter the room air. The half-life indicates how long it takes until the aerosol concentration at the place of measurement has decreased to half. The mean residence time characterizes how long the aerosols emitted at the respective measuring positions statistically remain in the room until they are separated by the sterilization unit. Table 1 shows the quantities determined from the measured decay functions for the different configurations.

**Table 1:**
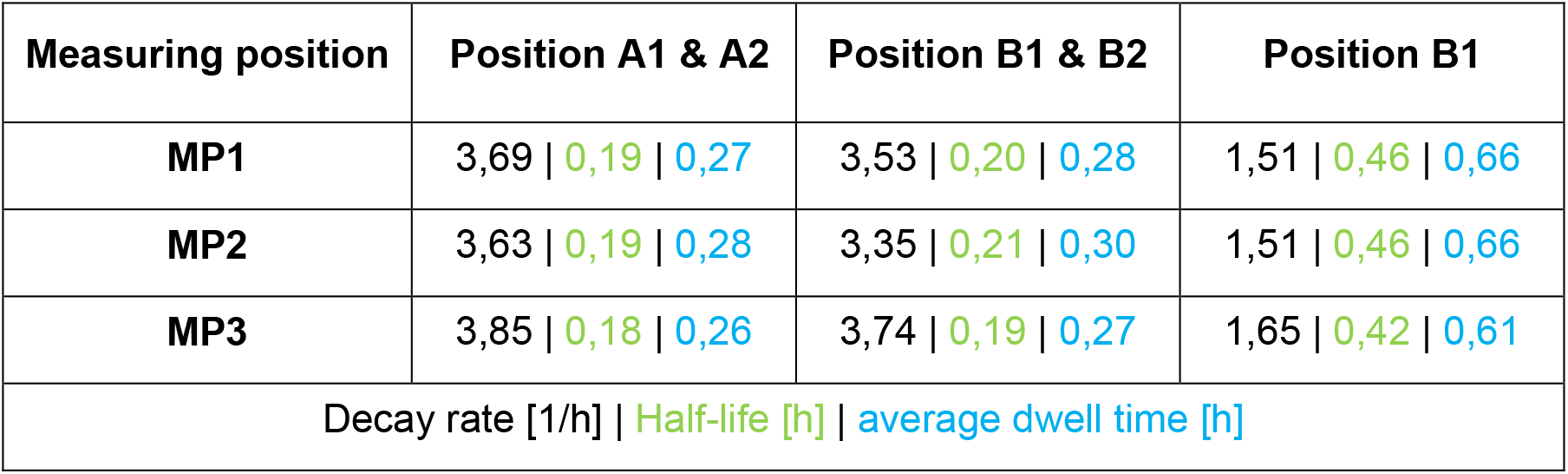
Decrease of aerosol concentration over time for different measuring positions and set-up configurations. Decay rate (black), half-life (green) and mean residence time (blue).

First of all, it can be stated that, despite the size of the room, the filter performance is only slightly dependent on the measuring position. The assumption that aerosol particles may remain in the corners of the room for a long time is therefore not justified. The turbulence in the room air is the reason why the concentration in the room is very uniform. Furthermore, the table shows that the location of the units does not have a significant influence on the filter performance. At 330 m^3^/h for each unit, the aerosol concentration is halved in about 11.5 minutes for configuration A1 + A2 and after about 12 minutes for configuration B1 + B2. According to this analysis, the influence of positioning on the filter performance is not very large. However, if only one disinfection device is used, the half-life is extended to about 25 minutes (MP3). This result clearly shows that the volume flow of the device must be very well adapted to the size of the room in order to achieve the desired filter performance. Small devices with a low volume flow rate cannot provide any protection against indirect SARS-CoV-2 infection in a large room. If the volume of the room is x m^3^, then the volume flow of the unit should be at least 6x/h to ensure sufficient security against a SARS-CoV-2 infection. For a room volume of about 200 m^3^, a volume flow of 1200 m^3^/h would be necessary to meet the requirement.

Due to the hazardous nature of SARS-CoV-2, this requirement of 6x/h should never be undercut. Statements that an air exchange rate of 1 − 2 is sufficient to effectively prevent SARS-CoV-2 infections are based on false assumptions. In areas where there is evidence of infected persons, air exchange rates of 12 − 15 are usually required [15, 16, 17]. An air exchange rate of 6 represents a reasonable and realistic compromise between safety and feasibility. If, despite an air exchange rate of 6 per hour, SARS-CoV-2 infections indoors cannot be effectively prevented, this could mean that a large number of infected persons were present in the room. An increase in the air exchange rate would then be advisable for the future.

However, the current infection figures suggest that this case is currently very unlikely. It could also be that the persons present have not been infected by an indirect infection due to the viral load in the room, but rather by direct infections that can occur when non-infected persons are coughed on by infected persons or when these persons talk for a longer period of time without sufficient distance. Disinfection devices and room air cleaners can do little against the direct danger of infection. The prevention of direct infections requires distances or the wearing of particle-filtering respiratory masks, mouth-nose covers, face shields or barriers made of Plexiglas.

The tested Viromed Klinik Akut V 500 disinfection device can be used in rooms up to 22 m^2^ in size, if the filter performance alone is considered. According to the manufacturer, however, the integrated ionization technology makes it possible to operate the unit in larger rooms of up to 50 m^2^ [20]. According to the manufacturer, the ionization of the air already largely inactivates the viruses at the point of release. According to the manufacturer, the device therefore not only offers protection against indirect infection, but also against direct infection in some cases. How effective the ionization is cannot be answered on the basis of the experiments carried out here, since no experiments with infectious viruses can be carried out in the laboratories of the institute. However, even without this protective function provided by ionization and taking the room size mentioned above into account, the device is powerful enough to significantly reduce the indirect risk of infection in treatment rooms, waiting rooms, normal offices, in the reception area of medical practices, in pharmacies or in dining rooms and lounges in old people’s homes, small stores, etc..

## 6. Dependence of the filter performance on the room geometry

The filter performance depends not only on the number of units and their location, but also on the geometry of the room. Especially in long rooms, aerosol particles can generally be separated less efficiently, since according to textbook opinion [12] the wall jet on the ceiling will eventually detach and a recirculation area will form that does not reach the opposite wall. This situation is comparable to the situation shown in Figure 4 (right), where the detachment of the flow from the ceiling in a long room is not caused by an object but by the reduction of the momentum of the wall jet with increasing distance. The reduction of the impulse is caused by the wall friction, the turbulent air movement and the entrainment visible in figure 7. The entrainment accelerates slow flow areas with aerosol particles by the fast wall jet and the work required for this leads to a reduction of the wall jet impulse. The turbulence primarily leads to a beam widening, which in turn leads to a local impulse decrease and therefore shifts the release position of the wall jet closer to the device. Due to these effects it seems plausible that the front area of the room is filtered very well, while the rear area of the room remains rather unaffected. In a previous work, however, it was already shown that this textbook opinion does not necessarily correspond to reality [8]. To investigate this situation with the Viromed Klinik Akut V 500 disinfection unit, measurements were taken in an elongated room with a cross-sectional area of approx. 4 m^2^. Two different room lengths were investigated: 22.4 m (see Figure 9 for experimental setup) and 11.8 m (Figure 10).

**Figure 9:**
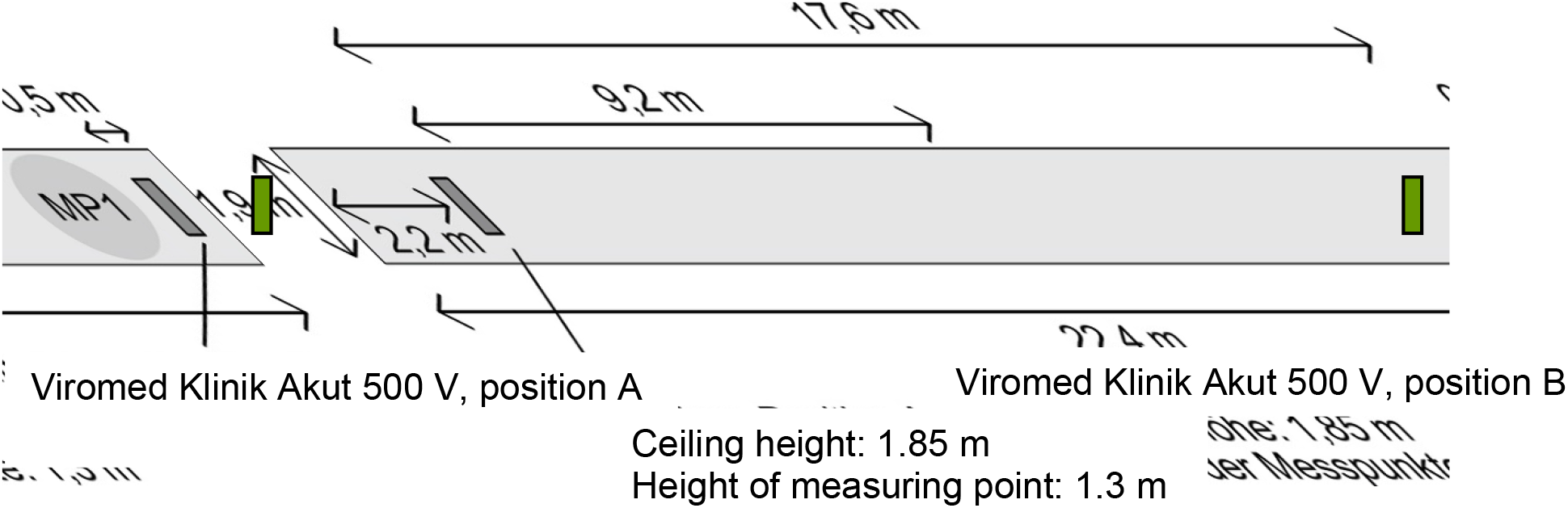
Arrangement of the components in the long corridor configuration for concentration measurements

**Figure 10:**
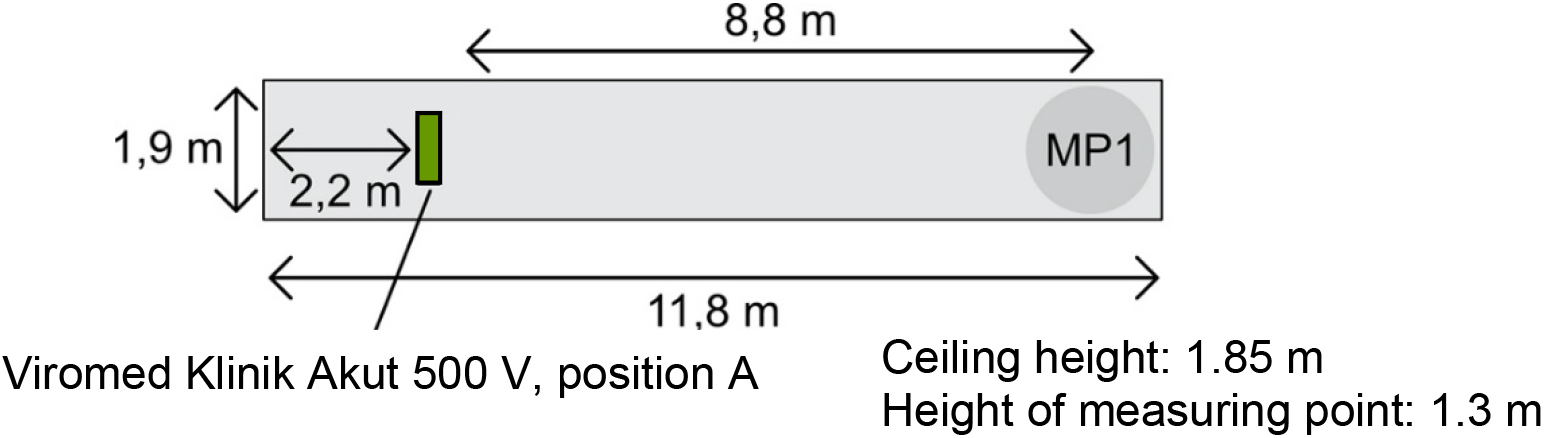
Arrangement of the components in the short corridor configuration for concentration measurements

The values determined from the concentration measurements in Figure 8 (right) are summarized in Tables 2 and 3 for the respective room sizes. It is clear that even in elongated rooms a rather fast filtering of the aerosol particles is achieved. If two units are positioned at the respective end faces of the room, the half-life is less than 5 minutes. If only one device is used, the half-life is 8 ½ minutes. Even at the distant measuring point MP2 in the long configuration, a significant decrease in aerosol particles is observed over time, which corresponds approximately to the decrease at the position MP1. The results show that in very long rooms, the use of two disinfection units at each end may be recommended when a very rapid decrease in viral load is required, e.g. in the corridor of a ward in a hospital/retirement home, a practice or in a hotel.

**Table 2:**
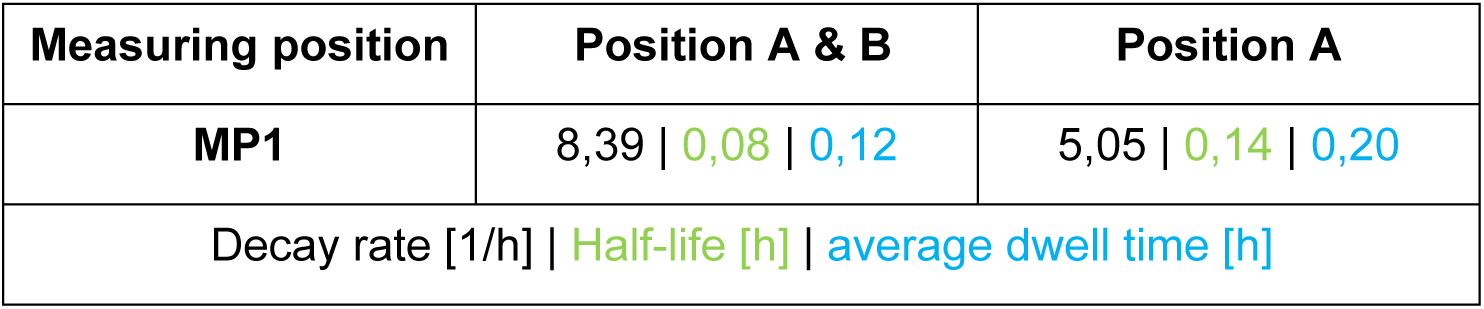
Decrease in aerosol concentration in the long corridor configuration over time for different configurations. Decay rate (black), half-life (green) and mean residence time (blue).

| Measuring position | Position A & B | Position A |
| --- | --- | --- |
| MP1 | 8,39 \| 0,08 \| 0,12 | 5,05 \| 0,14 \| 0,20 |
| Decay rate [1/h] \| Half-life [h] \| average dwell time [h] |  |  |

**Table 3:**
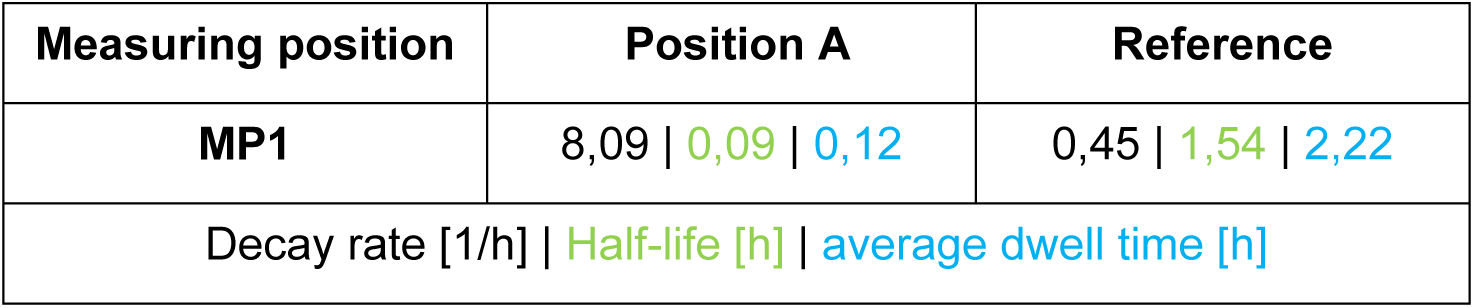
Decrease of aerosol concentration in the short corridor configuration over time. Decay rate (black), half-life (green) and mean residence time (blue).

Schematically, the temporally averaged flow situation in the long room can be shown with two devices as sketched in figure 11. As expected, the aerosol particles are filtered much faster in the short corridor configuration. According to table 3 the half-life is about 5 minutes at a volume flow rate of 330m^3^/h. The decay constant is 8.09, which means that at this room volume an air exchange rate of about 8 per hour is achieved. Without operation of the disinfection unit, the half-life is over 90 minutes. This comparison illustrates the potential for protecting people in the room by using disinfection units and room air cleaners in closed rooms.

**Figure 11:**
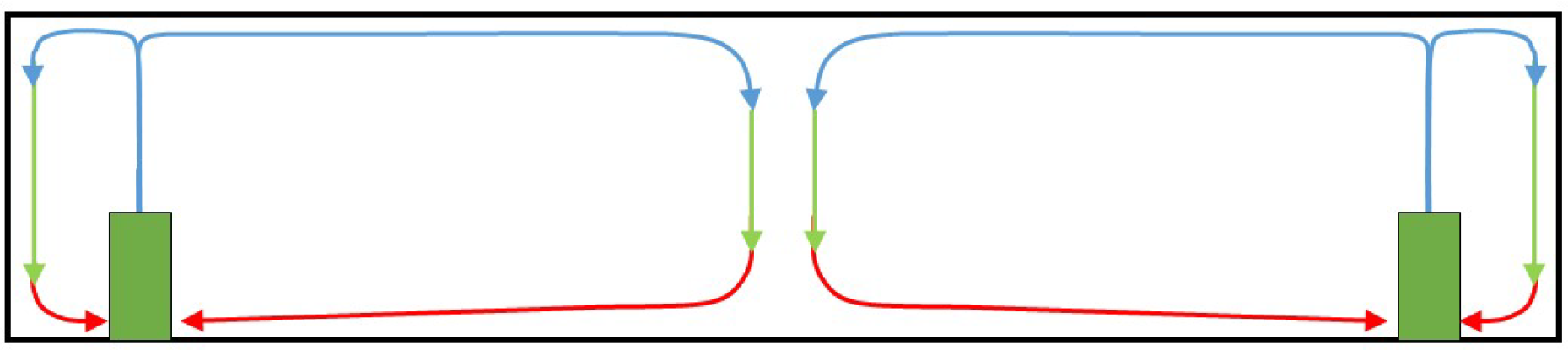
Idealized representation of the air flow in a long, empty room in which two disinfection units are operated. In reality, the flow phenomena are three-dimensional.

## Summary and Conclusion

The quantitative measurement results show that with the tested Viromed Klinik Akut V 500 disinfection device, the aerosol concentrations in a 22.5 m^2^ room can be halved in around 5 minutes. The air exchange rate in these cases is 8 per hour. This means that a single Viromed Klinik Akut V 500 disinfection device is very well suited to ensure a high level of safety from indirect SARS-CoV-2 infection in rooms up to approx. 30 m^2^. The room size corresponds to typical treatment rooms, waiting rooms, reception areas, pharmacies, offices,…

In a 42.5 m^2^ room, a half-life of 4.8 minutes was achieved with simultaneous operation of two Viromed Klinik Akut V 500 disinfection units, and the air exchange rate was 8.4. A comparison with the results in the smaller room clearly shows that scaling is easily possible. This means that if the room size is doubled, the air exchange rate, half-life and dwell time of the aerosol particles in the room do not change significantly, even if the number of units or the volume flow is doubled.

In rooms with 80 m^2^ and the operation of two Viromed Klinik Akut V 500 disinfection units, a halving of the aerosol particle concentration is achieved in approx. 11 minutes. The air exchange rate is therefore slightly lower than 4 per hour and thus below the value of 6 room air changes per hour recommended by us. However, since the device has an ionization unit in addition to the filtering of the aerosol particles, whose power can be freely adjusted on the device, an additional protection against a SARS-CoV-2 infection can be realized with this room size by ionizing the room air according to the product sheet of the manufacturer.

In even larger rooms, rooms with many large objects, ceiling interruptions due to beams or room-dividing lamps, or very angled rooms, sufficient disinfection units should be used to filter all areas quickly. Due to the danger of SARS-CoV-2 infection, the air exchange rate of the devices should not fall below a value of 6 air changes per hour, which is a good compromise between technical feasibility and safety from indirect infection. If additional protective mechanisms are available (ionization of the room air, UV-C, ventilation systems, free ventilation), they will have a supportable effect.

The results of the study show that powerful disinfection units can quickly reduce the aerosol concentration in rooms and keep it at a low level. Therefore, the indirect risk of infection can be reduced by these devices even with closed windows and without a suitable HVAC system. They are therefore very well suited to permanently ensure a low virus load in rooms such as treatment rooms, waiting rooms, reception areas, pharmacies, old people’s homes, offices and stores. The regular opening of windows is not necessary and the well-being in the room is not affected. They also offer the advantage over HVAC systems that are operated with little or no fresh air, that the viruses are really separated and inactivated by means of UV-C radiation and ionized charge and are not distributed through other channels in the building. In addition, the units are energy-efficient, since the costly heated room air is not led outside, as in the case of free ventilation or HVAC systems with a high proportion of fresh air, but only the harmful components of the room air (viruses, bacteria, pollen, fine dust, …) are separated and inactivated. Thus, these devices not only contribute to the improvement of the indoor air quality, but also to climate protection during the pandemic.

When purchasing disinfection devices or room air purifiers, it is very important that the devices also have reliable equipment for separating and inactivating the viruses. Low-priced units usually have neither sufficiently large volume flows (6 times the room volume per hour!) nor efficient filters of class H13 / H14 or comparable, which provide the required degree of separation at the large volume flows, or powerful ionization or UV-C units.

In conclusion, it should be emphasized that although disinfection units are suitable tools for reducing the indirect risk of infection, they can only not prevent the direct risk of infection, which can be caused by direct coughing or during long maintenance over short distances, by ionizing the air. It is therefore important to maintain sufficient distance from other people, to wear simple mouth-nose covers or face shields or to protect oneself from direct SARS-CoV-2 infection by means of Plexiglas barriers.

## Data Availability

Data is stored on a server of the institute and may be available on request.

## Note

The investigations were financially supported by the company Viromed GmbH, Rellingen. The Viromed Klinik Akut V 500 disinfection equipment was provided by Viromed GmbH for the investigations. The investigations were carried out in accordance with good scientific practice. The support by the company Viromed GmbH has no effect on the results presented.

